# AMEBIASIS: AN ANALYSIS OF ITS DISTINCTIVE PRESENTATION IN THE CONTEXT OF JORDANIAN CASES

**DOI:** 10.1101/2023.11.01.23297932

**Authors:** Enas Al-khlifeh, Ahmad B. Hasanat

## Abstract

Amebiasis is a serious health problem, particularly in economically disadvantaged countries. However, unresolved questions remain regarding the clinical symptoms and laboratory results of amebiasis in Jordan, a developing country. Therefore, we assessed the prevalence and severity of amebiasis in a sizable population in the center of Jordan in relation to potential risk variables such as environment, age and sex of the patients, collecting data from the medical records of the patients treated at the Al-Hussein Hospital in Al-Salt city between 2020 and 2022. Clinical manifestations and laboratory findings, including fecal red blood cell and leukocyte counts, total and differential leukocyte counts by age, and other information were included in this report. This study showed that amebiasis is a common problem in Al-Salt city, accounting for 16.8% of all gastroenteritis cases. Amebiasis affects individuals of all sexes and ages. When comparing males of different ages to females of similar ages, both age and male sex showed an increased risk of amebiasis (P=0.014). Infants and toddlers accounted for more than one-quarter of the cases, with more than half of them diagnosed with watery diarrhea. Numerous patients of all age groups displayed age-related neutrophilic leukocytosis and atypical WBC counts. The findings of this study suggest that amebiasis is highly prevalent, affects a significant proportion of infants and toddlers, and causes severe clinical symptoms. This is the first study on Al-Salt city population with respect to multiple variables and demographic factors. These data are crucial for the decision-making of medical professionals. The epidemiology of amebiasis in Jordan must be determined, and preventive measures must be developed, especially for childcare at educational facilities.

## 1. INTRODUCTION

Parasitic gastroenteritis (GE) is a serious global health concern. Amebiasis is transmitted by the fecal-oral route (“CDC - DPDx - Amebiasis,” 2019). The transmission occurs when a parasitic cyst in the infectious fecal matter is consumed via food or water. The acid-resistant cyst upon entering the small intestine, excysts to produce the organism’s motile and invasive form, trophozoites, which travel to the large intestine, and colonize the colon lumen as commensal flora or infiltrate the colonic epithelium, causing inflammation and amoebic colonic ulcers (Yue et al., 2021). Proliferation of the parasite in the colon results in symptoms of GE, including abdominal pain, dysentery, or mucous soft stools. However, the majority of infected people show no overt symptoms. The infection becomes life threatening if *E. histolytica* becomes invasive and spreads extra-intestinally to form liver abscesses because excess hepatic iron creates an ideal milieu for *E. histolytica* proliferation in the liver (Salles et al., 2003).

Poor hand hygiene and contamination of water bodies and underground water with fecal matter are the main risk factors for amebiasis (“CDC - DPDx - Amebiasis,” 2019). Amebiasis can affect people of any age, including children (Haque et al., 2006; Hegazi et al., 2013). Male sex is a known risk factor for the development of an amoebic liver abscess, especially in young men, although amebiasis can afflict women as well (Ghosh et al., 2014). According to previous studies, fecal contamination during oral and anal sex with homosexual or bisexual men increases the risk of amebiasis (Hung et al., 2011). Patients with compromised immune systems are extremely vulnerable (Zanetti et al., 2021).

Past research conducted in Jordan has revealed that amebiasis is a prevalent infection. However, several questions remain unanswered. Along with uncertainties regarding the clinical and laboratory signs, the severity of amebiasis in Jordanian patients of all ages and sexes remains unknown. This study aimed to determine the prevalence of amebiasis in a significant population in Jordan’s central region in relation to potential risk factors to understand the epidemiological profile of amebiasis in Jordan. In addition, we used random forest machine learning algorithm to classify the important laboratory features and risk factors associated with the severity of infection

## 2. MATERIALS AND METHODS

### 2.1. DATA COLLECTION

Data were collected from Jordan’s Hakeem electronic health record system between January 20, 2021, and August 28, 2022. Since 2009, Hakeem has managed and collected medical data from Jordan’s public healthcare system (“Hakeem Program | Electronic Health Solutions,” n.d.). The data used for the analysis included wet-mount reports of the detection of intestinal parasites. Laboratory tests were performed in accordance with the normal operating procedures of the Jordanian health centers. Briefly, stool samples were collected in labeled, clean, dry, leak-proof, and sterile plastic containers for a laboratory diagnosis from all patients with gastroenteritis (GE) with suspected involvement of an intestinal parasite. Within 30 min of sample collection, direct stool examinations were performed to examine for the presence of common intestinal parasites, including *E. histolytica/dispar, Giardia lamblia*, and worms such as *Ascaris lumbricoides, Hymenolepis nana, Enterobius vermicularis, Strongyloides stercoralis*, and *Trichuris trichiura*, hookworms besides *Taenia* and other cestode species. After stool examinations were completed, the infected patients were treated according to the national guidelines.

Positive parasitology results for *E. histolytica* were exclusively included in this study because it is the predominant protozoan reported. A few sporadic cases of *Giardia* (less than 10) were reported during the study period but were not included in our analysis. Hakeem’s sociodemographic data and laboratory results were examined and compiled using a worksheet specifically designed for this purpose. The dataset contained the date of sample collection, sex, age (years), diagnosis (ordered categorical), and stool examination results with severity indicators such as stool-red (RBCs) and leukocytosis or white blood cell (WBCs) counts. The WBC count was categorized into three based on the number of cells/high field: low = 0 to 3, medium = 3 to 6, and high = more than 6. In addition, results of the complete blood count (CBC) were used to obtain WBCs and neutrophil counts as it is considered a sign of the reaction to the infection and severity. The patients were categorized into three age groups: toddlers (less than five years), school-going age (represents 5–18 years), active workforce (19–59 years old), and adults (older than 60 years).

### 2.2. DATA PROCESSING, ANALYSIS, AND VISUALIZATION

The data were curated after extraction from Hakeem using the Pandas Python package (2023). The python packages numpy (Harris et al., 2020), scipy.stats (Virtanen et al., 2020) and statsmodels (Seabold and Perktold, 2010) were used to categorize the data and establish the association between variables and display their distribution patterns. In order to compare qualitative data, a Chi-square test was performed, which clarified the distributions of categorical variables, specifically the relationship between risk factors for amebiasis, including sex, age, and other laboratory findings that indicated the severity of infection, including the severity of diarrhea or the presence of RBC or WBC in the stool. Correlations among age, WBC count, neutrophil count, and neutrophil percentage were tested using one-way analysis of variance. Student’s t-test was used to compare numerical variables (WBC count, neutrophil count, and neutrophil percentage) and to determine whether there was a significant difference in the mean between groups. The analysis adhered to a significance threshold, wherein a P-value of less than 0.05 was deemed statistically significant, ensuring the reliability and validity of the results. The code used for the analysis is included in Supplementary File (S1). Graphical data representations were generated using Matplotlib. pyplot package (Hunter, 2007) and Seaborn in Python (Waskom et al., 2017). We have also included a brief excerpt from the dataset in the Supplementary File (S2).

## RESULTS

We performed a retrospective study on patients’ parasitology analysis data from Al-Hussein/Salt Hospital in Jordan, collected between September 2020 and December 2022. To identify intestinal parasites in the study area, 4429 stool tests were performed over the study period on patients with GE symptoms. Among these, 763 were confirmed to have amebiasis (16.98%), with 381 male (49.9%) and 382 (50.1%) female. The study participants were between the ages of less than one month and 99 years. The median age of female patients was 24 years, whereas that of male patients was 19 yrs. There was no significant difference between the median ages of females and males (P = 0.43, Figure 2). The prevalence of amebiasis among males and females across various age groups was not significantly different (P = 0.39). However, among the toddlers and preschoolers, there were more males than females (Figure 2A). In contrast, there were considerably more females than males in the adult group (aged 19 to 59).

Among all age groups studied, adults aged 20–59 years had the highest infection rate, whereas those older than 60 years had the lowest infection rate (Figure 2A). The percentages of amoebic infections across age groups did not differ significantly (P= 0.06). According to the clinical signs, the intensity of the symptoms was as follows: abdominal pain without diarrhea, 145 (19%); semisoft stools, 189 (24.7%); watery diarrhea, 426 (55.8%); rectal bleeding, 3 (0.3%). Individuals aged 1 month to 4 years accounted for 27% of the total, with 107/206 cases (51.9%) diagnosed with watery diarrhea. As expected, WBC and neutrophil counts showed a correlation with the diagnosis. When patients were diagnosed with watery diarrhea, these measurements increased (Figure 3). The correlation between other laboratory features and the diagnosis is shown in Figure (2B).

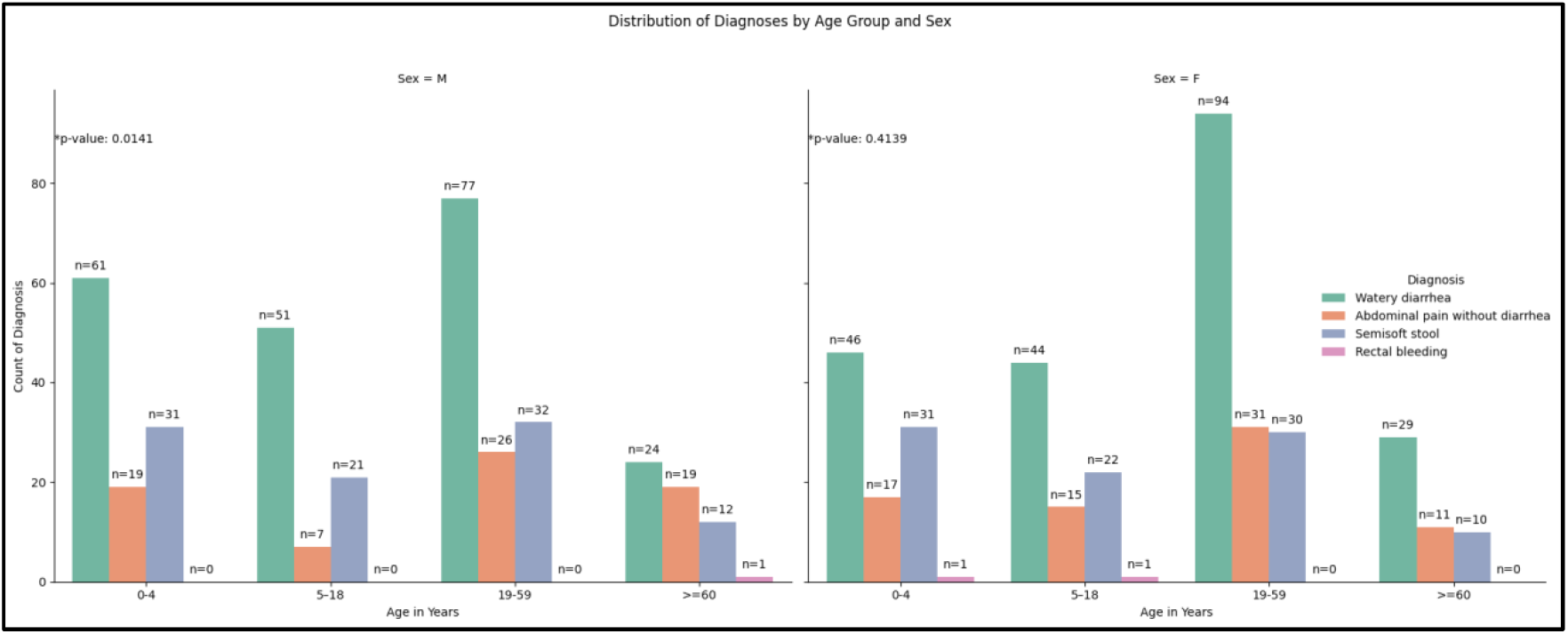

The following results were found when the risk variables and diarrhea severity were compared: each diagnosis had a different median age for males and females (Figure 1A). Both males and females under 48 years of age accounted for the majority (75%) of the recorded cases, but many older individuals had watery diarrhea.

**Figure 1.**
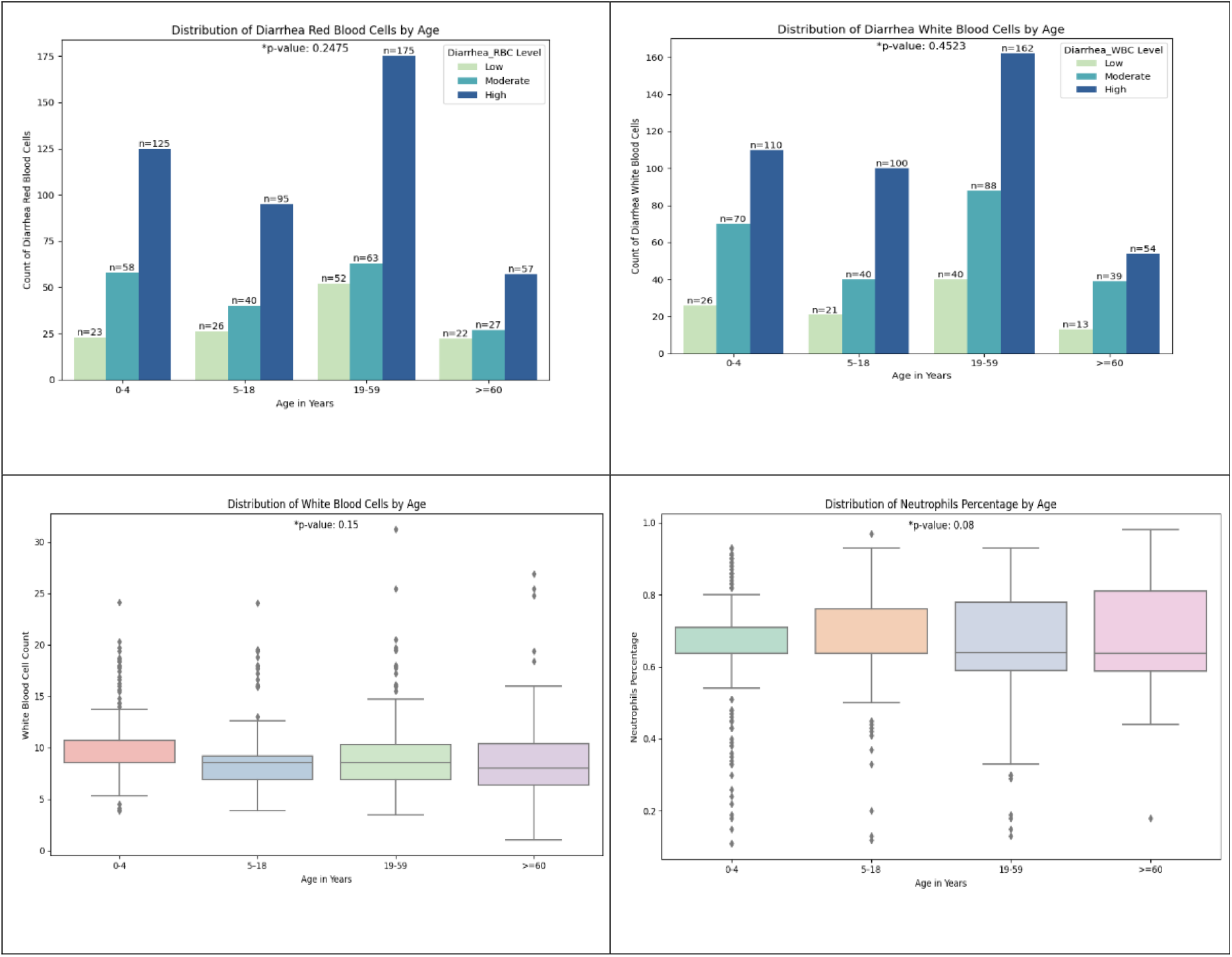
Clinical signs and laboratory findings with respect to age among patients with confirmed case of of *E. histolytica* gastroenteritis. P value < 0.05 indicates a significant difference. *n*= 763.

**Figure 2.**
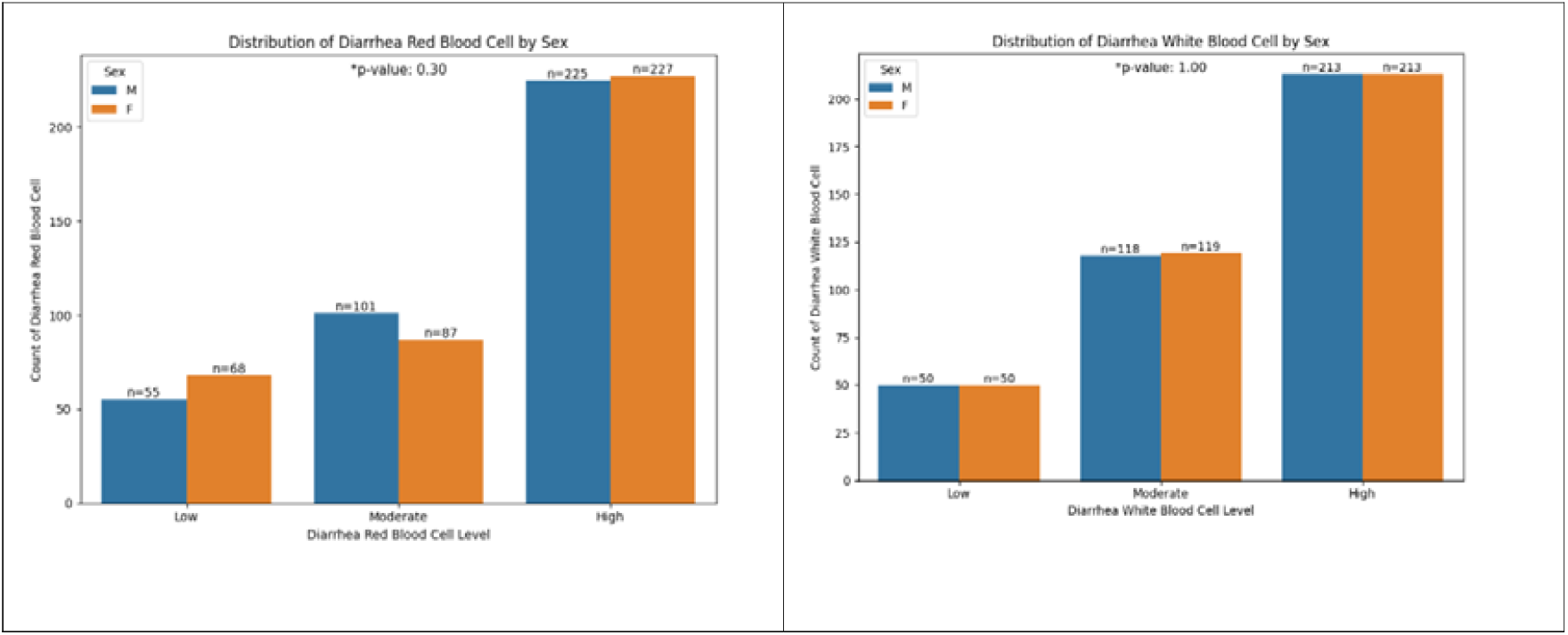
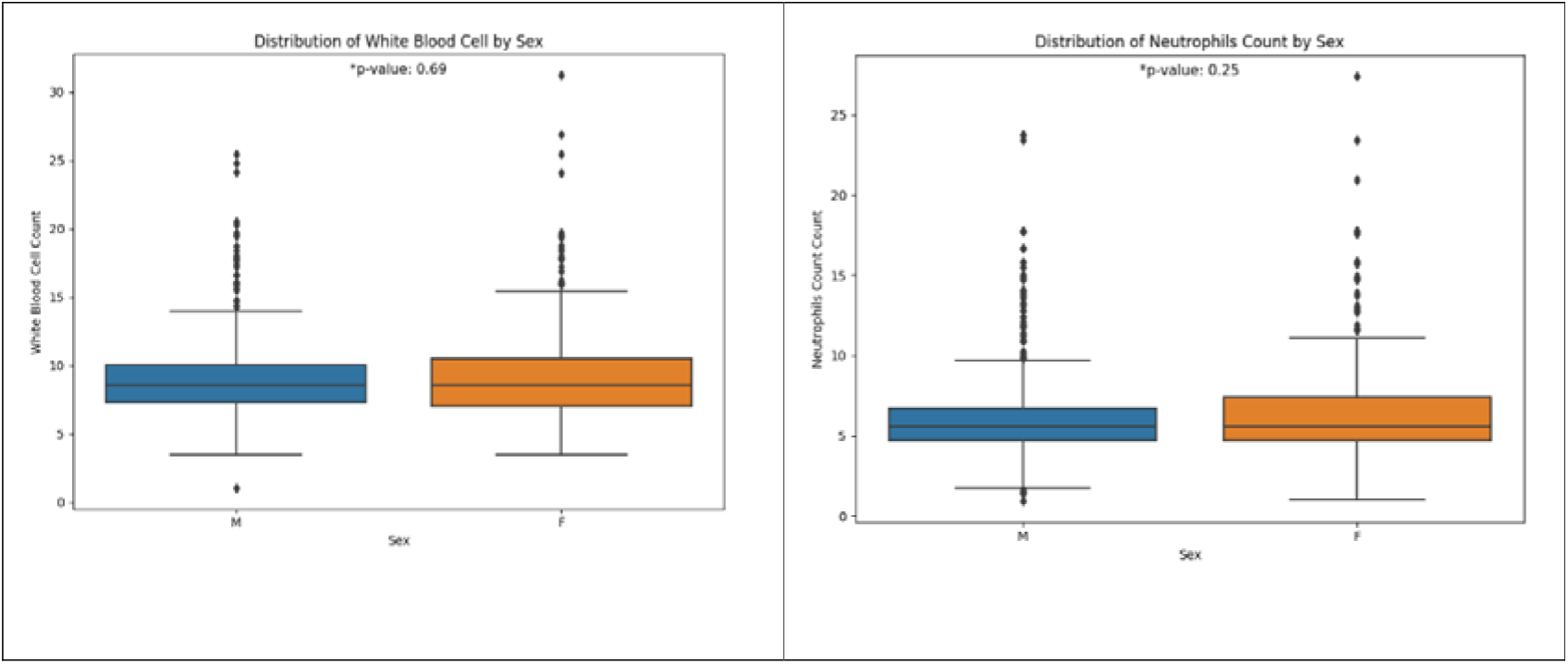
Clinical signs and laboratory findings with respect to sex among patients with confirmed case of of *E. histolytica* gastroenteritis. P value < 0.05 indicates a significant difference. *n*= 763.

There were no significant variations in the diagnosis of loose stools according to age group. However, a significant difference was found in males according to age when sex, age group, and diagnosis were analyzed (P=0.014; Figure 1). All severity features, including the stool-RBC, stool-WBC, WBC, and neutrophil count for the age and sex groups, were statistically insignificant (P ≥ 0.05) (Figures 2).

## 3. DISCUSSION

During the study, 4429 cases of GE diagnoses and requests for stool tests were recorded in the main hospitals of Al-Salt city. A total of 763 cases had confirmed amebiasis diagnoses, accounting for 16.8% of all GE cases. This demonstrates that GE-associated amoebiasis is a common public health concern in Al-Salt city.

The primary technique for verifying the presence of amoebiasis was conventional microscopic analysis of stool samples. Compared with other detection methods, such as the *E. histolytica* antigen detection test, it has been shown to have higher sensitivity and specificity for identification (Al-Dalabeeh et al., 2020). However, according to our assessment of computerized data, confirmed instances of amoebiasis displayed GE symptoms, including vomiting, abdominal pain, diarrhea, and bloody stools. There are seven species of Entamoeba that can infect people, *E. histolytica* is commonly acknowledged as a disease-causing organism. Whereas *E. dispar*, which is morphologically identical to *E. histolytica*, is mostly non-pathogenic and associated with asymptomatic infection. At present, it is advised to use molecular techniques to precisely differentiate between pathogenic species of Entamoeba. Therefore, the confirmation of the exact species responsible of amoebiasis is outside the scope of this study, since we relied on retrospective analysis of lab test results.

An overall prevalence of 0.1–80.7% for amebiasis has been reported in the majority of earlier studies conducted in different regions of Jordan (Nawafleh et al., 2014; Abdel-Dayem et al., 2014). Although this claim was supported by the results of molecular typing and serology, the prevalence of the pathogenic type *E. histolytica* might be overestimated when using microscopy (Hijjawi et al., 2021; Al-Dalabeeh et al., 2020). There were disparities in the prevalence of infection between our findings and earlier reports from other Jordanian locations. A diagnosis is traditionally made based on the evaluation of a spherical cyst (∼10 mm) or trophozoit and is considered to be subjective. Additionally, geographical elements such as the source and nature of drinking water may have an impact on the presence of variance between various locales.

Demographic factors such as age and male sex pre-disposed to amebiasis infection. Males appear to acquire amebiasis at an earlier age than females. Our results agree with those of the previous Jordanian (Nawafleh et al., 2014) and global studies (Nasrallah et al., 2022). Collectively, these studies demonstrate a correlation between age and male sex with amebiasis. This bias is mostly due to high-risk sexual behaviors (Hung et al., 2011) or differences in immunological responses to infection between the sexes. For example, men have immunological characteristics, such as sex chromosomal complement, immune cell receptors, and sex hormones that make them more vulnerable to serious infections (Jacobsen and Klein, 2021; Lotter et al., 2013; Bernin et al., 2014).

Young children and toddlers under the age of five were discovered to make up more than a quarter of the patients in this study, with more than half of them having been diagnosed with watery diarrhea, which indicates a high severity of infection, as well as the presence of a significant proportion of aberrant WBC and neutrophil counts for the age. This could have severe effects on children of this age as diarrheal infections linked to amebiasis are more likely to cause dehydration, stunted growth, and induce malnutrition (Mondal et al., 2006). Severe complications caused by dehydration necessitate immediate hospitalization. The significant prevalence of amebiasis that we discovered in this study is higher than what was 20 years ago for Jordanian children of the same age group (Youssef et al., 2000). Furthermore, amebiasis in infants younger than one year was more prevalent than predicted. This may be considered an unusual manifestation of amebiasis at this age, given that the transmission of intestinal parasites is frequently associated with the contamination of water and food and that infants are not normally expected to develop amebiasis. However, the risk of infection is increased by inadequate breastfeeding, mixed feeding, and daycare (Hegazi et al., 2013). This finding emphasizes the importance of carefully observing the GE infection dynamics in Jordanian children to ascertain whether the increasing frequency of amebiasis in children is also prevalent in other Jordanian regions. There have been a few occasional reports with inconsistent results. Specifically, a prior surveillance research conducted in the capital city of Jordan discovered that the frequency of intestinal parasites (including *E. histolytica*) was the highest in children under the age of five (Chazal and Adi, 2007). Additionally, children under the age of 15 make up more than 60% of affected patients in northern Jordanian cities (Jaran, 2017). In contrast, adult patients in southern sites have *E. histolytica* at higher rates than children (Nawafleh et al., 2014). It is essential to obtain more information on the socioeconomic situation, water quality, dietary background, nursery, and daycare, to understand the causes of amebiasis in Jordan’s toddlers and young children.

amebiasis is a disease with varying degrees of severity because it frequently causes silent and chronic infections in contagious patients. In this study, some patients were diagnosed with abdominal pain without diarrhea. It has been proposed that Entamoeba sp. including *E. histolytica* can survive, reproduce, and disseminate without virulence (Marie and Petri, 2014). The parasite may, however, have genetically distinct isolates. One of these genotypes may have caused the severe manifestations observed in the studied cases. *E. histolytica* genotyping falls beyond the scope of this study. The host’s immune response, history of infection, and previous exposure may have an impact on the severity of infection in addition to the genotype of the parasite. Thus, further research is required to pinpoint the underlying genetic and immunological mechanisms responsible for the aggressive presentation of amebiasis in our study.

## Data Availability

Data will be available on request.

## LIMITATIONS OF THE STUDY

This study had some limitations. This study was performed in the Jordanian city of Al-Salt and was primarily focused on a single community. A more comprehensive, multicentric study involving different cohorts of patients from diverse ethnic, socio-economic, and regional backgrounds would provide more concrete data and reproducible patterns on the distribution of potentially diverse genotypes of the parasite and respective differences in the pathogenesis and immune responses upon infection. However, a sufficient sample of the local population was reflected in the number of patients admitted to city-only hospitals. Investigation of the immunological characteristics such as cytokine measurements and genotyping of the isolated Entamoeba species would have been beneficial in elucidating the potential mechanisms underlying the severity of amebiasis in Jordanian patients.

## CONCLUSIONS AND RECOMMENDATIONS

Amebiasis was found to be highly prevalent among patients with GE across all age and sex groups in the population under investigation. However, younger men were susceptible to amebiasis and exhibited severe clinical symptoms such as diarrhea. Typical clinical manifestations and laboratory findings of invasive amebiasis, such as significant WBC counts and neutrophilic leukocytosis, were observed in all age and sex groups. As per our findings, amebiasis can have a very high chance of recurrence depending upon the environmental and host variables primarily linked to sex, inadequate childcare, and irregularities in the water supply. As discussed before, the epidemiology of amebiasis in Jordan must be determined on a large scale, and preventive measures must be devised to curb the incidence in high-risk areas such as childcare and educational facilities. Additionally, to lower the prevalence of infection with amebiasis and other infectious GE agents in Jordan, periodic inspections should be performed during the transport and distribution of commercial water because the majority of Jordanians use it.

## ACKNOWLEDGMENTS

We acknowledge Jordan’s AL-Hussein/Salt Hospital for allowing access to medical records from the microbiology laboratory section. In addition, we gratefully acknowledge Mrs. Qamar Al-Mimi Geographic Information System Specialist from Nature Conservation Monitoring Center for providing a map of the study area.

## ETHICAL APPROVAL

This study was approved by the scientific and administration committee of scientific research at Al-Balqa applied university and Jordan’s AL-Hussein/Salt Hospital.

### Approval number

ID: 35/4/1205. The material is the authors’ own original work, which has not been previously published elsewhere.

## DATA AVAILABILITY

The dataset is constantly being updated, and a subset can be made available on request.

## SUPPLEMENTARY MATERIALS

Supplementary material associated with this article can be found, in the online version, at doi:

