## Supplementary figures and images for "AMEBIASIS: AN ANALYSIS OF ITS DISTINCTIVE PRESENTATION IN THE CONTEXT OF JORDANIAN CASES"

### Supplemental Table 1

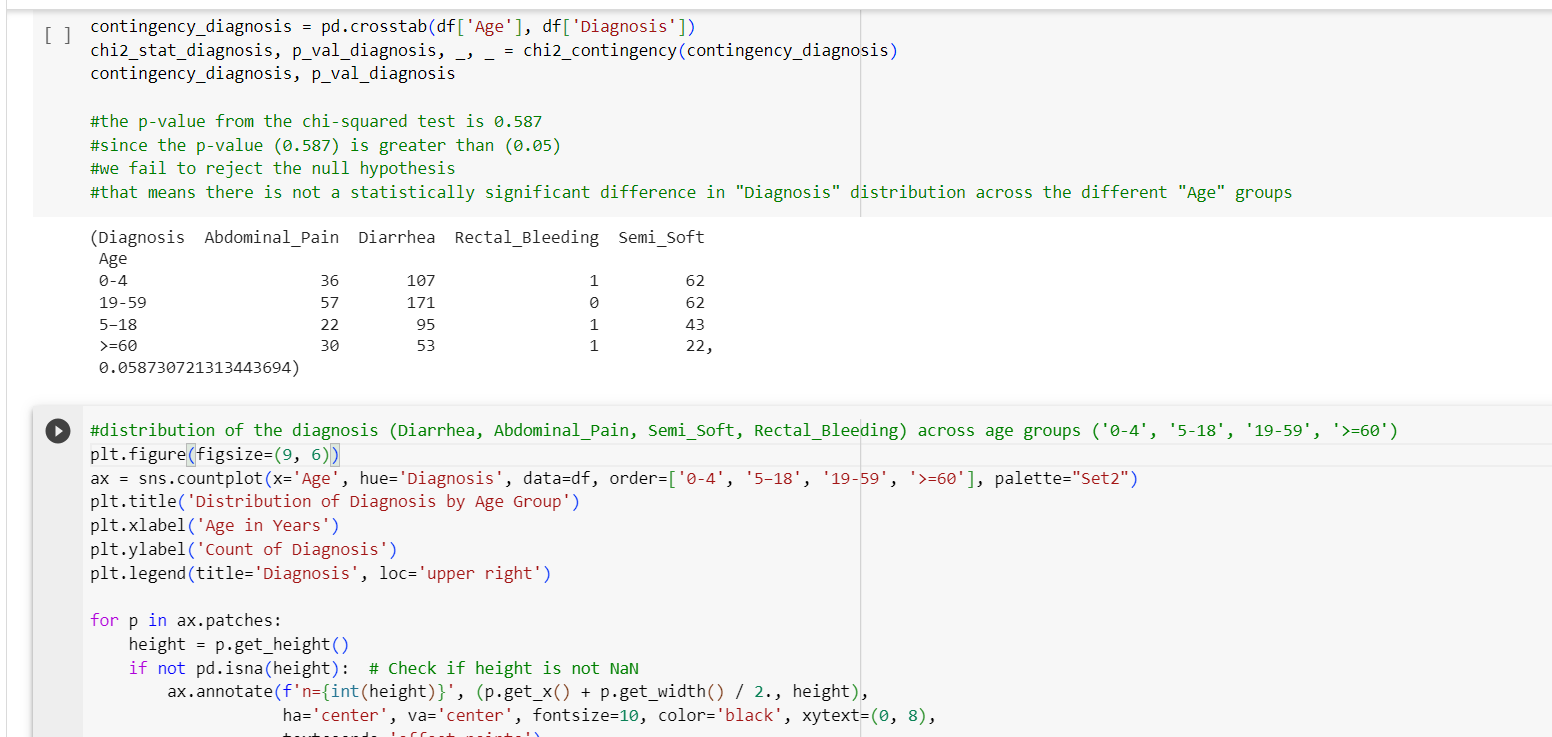

### Supplemental Table 2

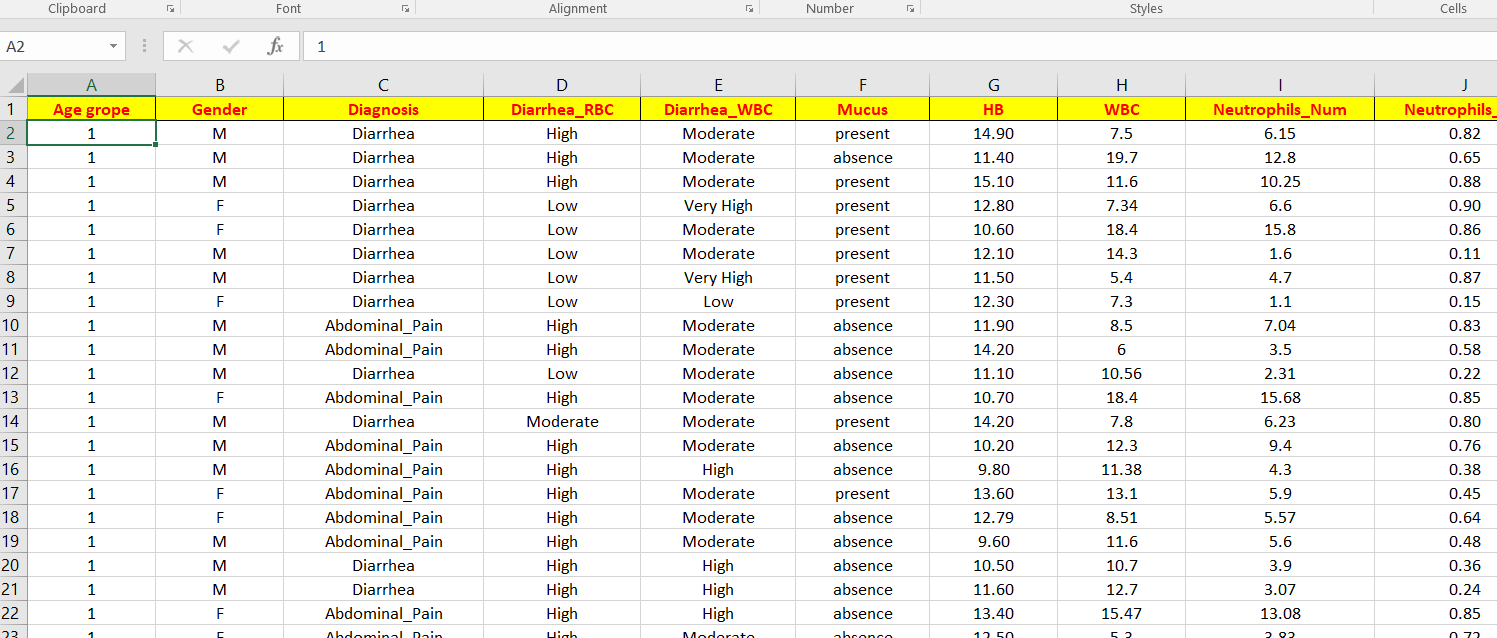
